# Deep learning-based physical exercise assessment of older adults using single-camera videos

**DOI:** 10.1101/2025.08.30.25334353

**Authors:** Vayalet Stefanova, Evelien Maeyens, Jeroen Brughmans, Dimitri Vargemidis, Julien Lebleu, Leen Stulens, Sanne Broeder, Karen Gilis, Mieke Deschodt, Benedicte Vanwanseele, David Beckwée, Moran Gilat, Benjamin Filtjens, Bart Vanrumste

**Affiliations:** Department of Electrical Engineering (ESAT), KU Leuven, Leuven 3001, Belgium; Faculty of Engineering Technology, KU Leuven, Leuven, 3000, Belgium; Department of Computer Science, KU Leuven, Leuven, 3001, Belgium; moveUP, Ghent, 9000, Belgium; in4care, Leuven, 3000, Belgium; Zorg Leuven, Leuven, 3000, Belgium; Department of Public Health and Primary Care, KU Leuven, Leuven, 3000, Belgium; Department of Movement Sciences, KU Leuven, Leuven, 3001, Belgium; Rehabilitation Research Group, Vrije Universiteit Brussel, Jette, 1090, Belgium; Neurorehabilitation Research Group (eNRGy), Department of Rehabilitation Sciences, KU Leuven, Leuven, 3001, Belgium; Department of Engineering Systems and Services, Delft University of Technology, Delft, The Netherlands; Institute for Health Systems Science, Delft University of Technology, Delft, The Netherlands

**Keywords:** Deep learning, Exercise monitoring, Motion analysis, Older adults

## Abstract

**Background and aim:** Regular physical activity preserves functional independence in older adults, yet care-home residents often miss out because personalized supervision is scarce. Autonomous, technology-supported exercise platforms could deliver such guidance without additional staff time—but only if sessions are automatically monitored for safety and quality. We therefore designed a deep learning (DL) system that (a) recognizes individual exercise types and (b) estimates joint angle trajectories from a standard video recording. These outputs are used to compute objective exercise performance metrics (EPMs) such as duration, repetition count, motion variability, and range of motion.

**Methods:** Seven care-home residents (aged between 65–94 years) performed six common rehabilitation exercises in front of a single camera while wearing 17 inertial sensors (Xsens MVN Awinda) that provided ground-truth joint angles. Two-dimensional skeleton poses estimated from the video were fed into a temporal convolutional neural network to recognize the exercises and estimate three-dimensional joint angles. We evaluated exercise segmentation with F1@50 and angle regression with mean per-joint angular error (MPJAE) across nine trunk and lower-limb joints, using leave-one-subject-out cross-validation. Pearson correlations assessed agreement between estimated and ground-truth EPMs.

**Results:** The DL model achieved an F1@50 of 0.92 (***±*** 0.04) for exercise recognition and an MPJAE of 7.7° (***±*** 0.91) for joint angle estimation. The estimated EPMs aligned closely with ground truth, achieving correlation scores of 0.93 (95% CI [0.90, 0.95]) for duration, 0.86 (95% CI [0.80, 0.90]) for repetition count, and between 0.3 and 0.9 for motion variability and range of motion across exercises.

**Conclusion:** The DL algorithm reliably estimates key exercise outcomes from a single video stream. This video-based monitoring pipeline could enable unsupervised, technology-supported exercise assessment in residential care homes while safeguarding session quality and safety. Future work will validate the approach in larger and more diverse cohorts.

## 1 Introduction

The aging population is emerging as one of the leading socio-political challenges of the 21st century. According to demographic projections by the United Nations, the proportion of adults aged 65 and above is set to more than double from 9% in 2019 to 16% by 2050 [1]. With age there is a decline in physical and mental capacities, which diminishes independence among older adults [2, 3]. Physical inactivity and sedentary behaviour plays a major role in this decline, whereas in most cases regular exercise has been shown to help delay or prevent these negative effects [4, 5]. Regular physical exercise has also been associated with improved quality of life as it promotes healthy aging and disease prevention both for people living at home and in residential care facilities [6, 7].

However, maintaining adherence to prescribed exercise regimens remains a major challenge [8]. Healthcare providers often prescribe self-directed exercises as part of rehabilitation therapy, but patient adherence is frequently low, undermining the effectiveness of these interventions [9]. This issue is especially prevalent among older adults living in residential long-term care facilities, who typically do not engage in sufficient physical activity to meet health recommendations [8]. Caregivers try to encourage older adults to perform physical exercises through personal guidance [10]. Access to such guidance is, however, not universally available due to the decreasing number of caregivers, the growing number of residents, and high financial cost of personalized interventions [11].

To address this challenge, various technologies have been developed to promote physical activity. These include exergames, which combine physical exercise with game-like tasks to enhance motivation, enjoyment, and engagement [12, 13]. Virtual reality simulators [14] and dance programs [15, 16] offer a similar solution by introducing variety into routine exercise programs, potentially improving adherence and overall satisfaction. Humanoid robot coaches have also been evaluated in terms of their acceptance and ability to improve engagement by providing real-time feedback of upper body exercise performance [17–19]. The envisioned future of humanoid robotic exercise coaching systems is to autonomously track users’ 3D articulated movements, deliver personalized motivational and corrective feedback, and continuously monitor safety [20].

Beyond motivating older adults to move more, an important application of these technologies is to support caregivers in evaluating how well individuals adhere to prescribed exercise routines and how effective those routines are in achieving therapeutic goals [21, 22]. This aligns with the broader goal of monitoring key biomarkers of healthy aging, which, as suggested by the World Health Organization, can provide insights into an individual’s functional status and risk factors for decline [23]. Traditional monitoring methods often rely on specialized equipment such as inertial measurement units (IMUs), depth cameras, or multi-camera setups. These systems are sometimes used in combination, such as pairing IMUs with video cameras, to improve motion tracking, and may also be supplemented with additional physiological sensors for monitoring parameters like heart rate, muscle fatigue, or respiration [21, 24–26]. While such setups can yield rich multimodal data for assessing physical performance, they are typically impractical for continual use by older adults due to their cumbersome setup nature and the frequent need for caregiver assistance.

Standard single-camera video offers an accessible contactless solution for exercise monitoring which can be deployed on a range of platforms, including robotic coaches, as well as more affordable options such as mobile phones or simple webcams. Advances in computational methods, particularly deep learning [27], combined with extensive publicly available datasets [28], have enabled the development of pose estimation algorithms such as OpenPose and AlphaPose [29, 30]. These algorithms estimate 2D image-plane positions of anatomical points (e.g., ankles and knees) in each video frame.

In the field of computer vision, numerous methods have been proposed for monitoring physical activity based on 2D poses which exclude the depth dimension as input [31–35]. However, these algorithms are typically trained and validated on young participants and may exhibit biases, particularly when applied in the context of exercise monitoring of older adults who typically move slower and with less range of motion [36–38]. Existing video pose-based methods for older adults largely target isolated rehabilitation tasks, such as sit-to-stand (STS) [39, 40] and gait analysis [41, 42]. The clinical outcome metrics that are computed are then limited to the rehabilitation goals of the particular task such as STS phases and their duration [39], cadence, and step time [42]. Other approaches provide a single metric such as a mobility score for a standardized mobility test [43] or movement similarity score with a reference coach during exercise performance [44].

Importantly, existing methods do not offer comprehensive performance monitoring across a wide range of exercises involving both upper and lower body movements in older adults. To address this, we propose a deep learning (DL) framework capable of estimating a clinically meaningful set of exercise performance metrics (EPMs) to support accessible, longitudinal monitoring and the potential for personalizing exercise programs by caregivers. The EPMs that we selected to compute align with the FITT principle (frequency, intensity, time, type) which is a well established fitness guideline [45]. The EPMs include (a) repetition count, which captures how often movements are performed and reflects the frequency component; (b) exercise duration, which measures the time spent engaging in an activity and corresponds to the time component; (c) range of motion, which reflects joint mobility; and (d) motion variability, which evaluates movement quality and stability providing insight into potential fall risk [46]. The combination of motion quality metrics, repetition count and duration can inform the adjustment of exercise intensity. To enable the computation of these EPMs, our DL framework (a) performs fine-grained exercise detection in terms of frame-level classification labels and (b) estimates joint angles in terms of the 3 rotational degrees of freedom: flexion/extension, lateral and axial bending. This approach supports objective tracking of an older adult’s exercise performance without the need for caregiver supervision.

## 2 Methods

### 2.1 Data acquisition

#### 2.1.1 Subject inclusion criteria

We recruited seven participants (mean age: 82.4 ± 8.1 years, 3 females and 4 males) from a residential care home in Leuven, Belgium. The home’s physiotherapist assisted in identifying potential participants, excluding those recovering from recent fractures or other injuries that could impede exercise performance. Participants were included if they had a transferring score of 1 or 2 on the Katz Index of Independence in Activities of Daily Living (ADL), indicating their ability to safely perform exercises without requiring assistance from a caregiver [47]. The transferring score evaluates the individual’s ability to move from one place to another, such as from a bed to a chair. A score of 2 indicates that the individual is independent in transferring when using a walking aid and a score of 1 indicates that the individual is fully independent in transferring. Prior to the first study-related procedure, all participants signed a written informed consent form approved by the ethical committee of KU Leuven and the university hospital UZ Leuven (EC Research UZ/KU Leuven: S61015).

#### 2.1.2 Data collection protocol

Each participant carried out five or six exercise sessions. Each exercise session included six exercises that were deemed safe to carry out unsupervised by a physical therapist.

All exercises were selected from the Geriatric Activation Program Pellenberg (GAPP), a rehabilitation program designed for individuals over 65 years old [48]. Exercises were selected to include a wide variety of movements to test the limits of the deep learning framework. Each exercise was performed twice within each exercise session and the order of the exercises was randomized between sessions and participants.

The measurement setup and the six exercises are visualized in Figure 1. The setup included a chair with armrests and sturdy tables positioned close together for support. Exercise equipment, such as a cup, was taped to the ground to prevent slipping (1). A webcam (Rifaa WebCam) was placed at a sufficient distance to capture the entire measurement setup. Camera footage was recorded at 30 frames per second with a resolution of 1920 x 1080. Two trained graduate students manually annotated each frame of the videos to identify physical exercises and split them into more fine-grained actions (Table 1) which we refer to as the sub-exercise action labels in this paper. A senior researcher verified the labels for correctness.

**Table 1:** Overview of the exercises and corresponding labels used in the analysis of this study. Each of the 6 exercises described in Figure 1 is split into sub-exercise actions which are used in the analysis of this study instead of the complete exercises. This segmentation was done by identifying the individual actions that collectively form a complete exercise. The activity labeled as 0 includes all movements unrelated to the instructed exercises.

| Label | Sub-exercise action | Complete exercise |
| --- | --- | --- |
| 0 | Random movements | Random movements |
| 1 | Getting seated on chair | Sit-to-stand |
| 2 | Standing up from chair |  |
| 3 | Bending forward while seated on chair | Bending forward/backward on chair |
| 4 | Leaning backward while seated on chair |  |
| 5 | Moving a weight from left to right armrest of chair | Moving a weight between armrests |
| 6 | Moving a weight from right to left armrest of chair |  |
| 7 | Lateral moving of the right leg | Lateral leg movement |
| 8 | Lateral moving of the left leg |  |
| 9 | Picking up a box and placing it on an elevated plank | Picking up a box from table/plank |
| 10 | Picking up a box from elevated plank and placing it in front of them |  |
| 11 | Moving right foot forward to touch a cup | Touching a cup with foot |
| 12 | Moving right foot backward after touching a cup |  |
| 13 | Moving left foot forward to touch a cup |  |
| 14 | Moving left foot backward after touching a cup |  |

**Fig. 1:**
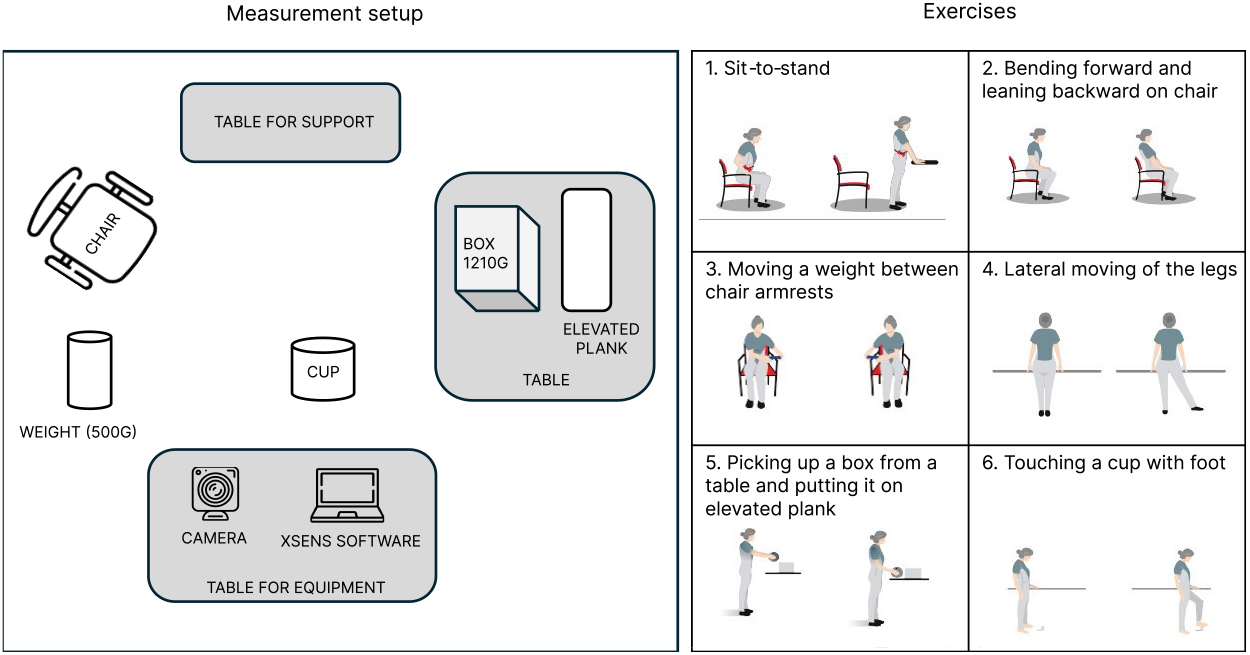
The data collection setup for exercise monitoring. **Left:** The setup includes a table for support, a main table with an elevated plank and a box (1210 g) for exercises, a table for equipment, which holds a camera and a laptop with a signal receiver and the Xsens MVN Analyze Pro software [49]. A chair with armrests and backrest is positioned for seated exercises, and additional tools like a cylindrical weight (500 g) and a cup are used for specific tasks. **Right:** The 6 physical exercises performed by participants during data collection.

We collected inertial-based motion capture (MoCap) data using Xsens MVN Analyze Pro (Xsens Technologies B.V., Netherlands) to use as ground truth for our regression task of estimating joint angles [49]. The MoCap system consisted of 17 wireless motion sensors (MTw Awinda) operating at 60 Hz, which estimated 3D joint kinematics. These 3D joint kinematics have been validated against gold-standard multi-camera marker-based MoCap systems [50]. Synchronization between the camera footage and motion sensor data was provided by the MVN Analize software through a LAN connection to the webcam. Prior to collecting data, the MTw sensors were calibrated using MVN Analyze Pro software (version 2022.01). This process appeared challenging for the older adults as some participants exhibited hand tremors during the phase requiring them to stand still while others had limited arm swing during the walking phase that differed from the expected healthy adult movement pattern. Consequently, this led to increased noise in the upper body joint kinematics. As a result, certain upper body joints were excluded from the analysis (see the Methods section for details). This highlights the need for caution when using IMU systems that rely on calibration based on movement patterns of healthy adults.

#### 2.1.3 Body keypoints extraction

The video recordings were first processed to estimate sequences of 2D keypoints. The use of 2D skeleton sequences has been shown to be effective for various downstream motion analysis tasks, providing a robust and reliable foundation for modeling motion across diverse datasets [51–53]. This approach has also been shown to work well with older adult populations [54], which motivated our decision to adopt it in this study. To extract 2D keypoints, we analyzed video segments where only the participant was visible, using AlphaPose, a state-of-the-art pose estimation algorithm [30]. AlphaPose provides high spatial resolution by detecting 17 anatomical keypoints for each video frame, including major joints such as the shoulders, elbows, hips, knees, and ankles. Each keypoint is represented by its *x* and *y* coordinates along with an associated confidence score indicating the reliability of the detection. Although AlphaPose processes frames independently and does not explicitly model temporal dependencies, the resulting frame-by-frame keypoint sequences implicitly capture the temporal progression of motion. This structure provides a frame-by-frame series of 17 *×* 3 matrices, ordered according to the COCO whole body keypoint format [55], which serves as input to our deep learning framework. All data was pseudonymized for privacy protection of the participants.

### 2.2 Deep learning framework

As shown in Figure 2, this study focused on the application of an existing fully convolutional architecture proposed by Pavllo et al. [31] to address two downstream tasks: (1) exercise recognition for frame-by-frame action annotation and (2) joint angle prediction for continuous kinematic analysis. The outputs of these tasks were used to compute the EPMs described in Section 2.3.3. The chosen DL model, referred to as VideoPose, is motivated by its state-of-the-art performance in the resgression task of 3D human pose estimation [31]. Moreover, it has also been successfully applied to classification tasks, such as detecting freezing of gait in patients with Parkinson’s disease and upper limb functional use in breast cancer survivors [56, 57]. Training strategies and hyperparameters followed the original studies of Pavllo et al. [31], and a testing procedure is outlined in Section 2.3.1. The implementation was carried out using Python version 3.12.2 and PyTorch version 2.2.1.

**Fig. 2:**
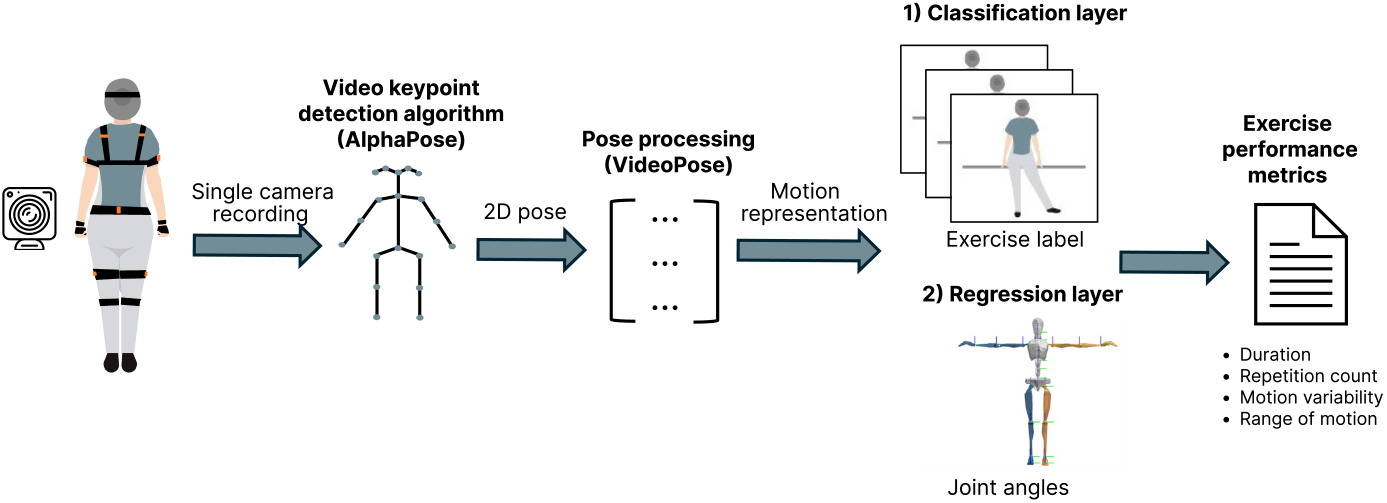
Overview of the workflow proposed in this study. Data is captured using a single camera and Xsens motion sensors. The AlphaPose algorithm extracts 2D poses from the video data, which are then processed by the VideoPose model to generate a motion representation. This representation is subsequently passed to a task-specific layer for either exercise recognition or joint angle prediction. A set of exercise metrics are computed based on the outputs of the two networks.

#### 2.2.1 Problem statement

The 2D pose sequence used as input to our framework can be represented as 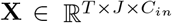, where *T* denotes the number of frames in the sequence and *J* is the number of joints (*J* = 17). The input channels are represented by *C*_*in*_ = 3, corresponding to the *x* and *y* coordinates and confidence score for each joint. Each sequence **X** is divided into overlapping windows of fixed length *k* and it is associated with a ground truth label vector **Y** which varies depending on the task. To generate predictions for each window *i*, the model learns a function *f*: **X**_*i*_ *→* **Ŷ**_*i*_ that maps the input window 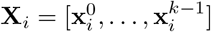 to an output **Ŷ**_*i*_ that is an approximation of the ground truth **Y**_*i*_ for that window. Each vector 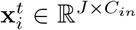 represents the 2D pose for frame *t* in the window. The task-specific details are outlined below:

- **Task 1:** The ground truth is a single action label corresponding to the central frame of each window, with *L* = 15 discrete action classes from the manual annotation of exercises. The model outputs a single label **Ŷ**_*i*_ *∈* ℝ^1*×L*^ per window of input data **X**_*i*_.
- **Task 2:** A sequence-to-sequence approach is implemented, where the ground truth corresponds to the 17 joint angles recorded with the motion sensors. Each keypoint is described by three angle types in terms of rotational degrees of freedom: *x* (lateral bending), *y* (flexion/extension), and *z* (axial rotation), which result in **Y**_*i*_ *∈* ℝ^*k×J×*3^.

#### 2.2.2 Deep learning network

The DL network implemented in this study was based on the VideoPose framework, which is a fully convolutional architecture proposed by Pavllo et al. [31]. This state-of-the-art model used temporal convolutions with ResNet-style blocks to capture temporal dependencies in 2D pose sequences. We followed the originally proposed pipeline, which outputs a 1 *×* 15 label prediction for Task 1 and 1 *×* 17 *×* 3 angle predictions for Task2, both generated per window of 2D pose data. All hyperparameters were retained from the original architecture, including convolutional layers with 1024 output channels and a dropout rate of 0.25 [31]. Training was performed for 60 epochs with a batch size of 32. Following the protocol proposed by [31], the model was optimized using the Amsgrad variant of the Adam optimizer [58], with an initial learning rate of 10^*−*3^ that decayed exponentially by a factor of *α* = 0.95 after each epoch. For the input data, the 2D pose data was split into overlapping windows of 243 frames with 50% overlap, aligning with the data characteristics that the VideoPose architecture was optimized for [31].

**Task 1:** To perform exercise recognition, the original regression model was adapted for classification by modifying the final convolutional layer to output exercise labels instead of continuous values and adjusting the output tensor shape to match classification targets. Cross-entropy loss was used to optimize the classification performance.

**Task 2:** For the regression task of estimating joint angles, we used a loss function that adapts the Mean Per-Joint Position Error (MPJPE) to angular space (MPJAE) by computing mean absolute differences between predicted and ground-truth joint angles across all joints and angular dimensions:

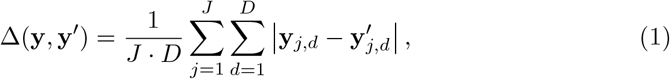

where **y**_*j,d*_ denotes the *d*-th rotational angle type (*D* = 3) of joint *j* in pose **y**, and *J* is the number of joints (*J* = 17). Angular differences were normalized to the range [0^*◦*^, 180^*◦*^] to account for circularity. Due to the reliability issues observed during sensor calibration and data collection, particularly noisy upper body joint kinematics, a subset of 9 joints was selected for this task. With this we ensured that only reliably recorded joint angle data was used as ground truth. A corresponding mask was applied to the loss function, focusing the training on the 3 axial joints (lumbosacral (L5S1), thoracolumbar (L1T12), and cervicothoracic (T1C7) [59]) and the lower body joints (left and right hip, knee, and ankle). This approach ensured that only the weights for reliable joints were updated, allowing the model to effectively learn the kinematics of clinically relevant angles.

### 2.3 Evaluation

To evaluate the performance of the models on the two tasks, a leave-one-subject-out (LOSO) cross-validation (CV) approach was implemented. In this method, the data is iteratively partitioned based on the number of subjects in the dataset. For each iteration, one subject is preserved for evaluation, while the data from the remaining subjects is used to train the model. This process is repeated until all subjects have been used for evaluation and the final results described for each of the two tasks represent the averaged performance across all iterations. The LOSO approach is particularly suitable for scenarios where models need to generalize well to new, unseen subjects, reflecting real-world applications [60] such as exercise assessment. However, the results are based on a small sample of seven participants, making this study more suitable as a proof-of-concept rather than for drawing broad conclusions.

#### 2.3.1 Experimental setting

To compute the EPMs, predictions at a frame-by-frame resolution were required. During training, overlapping input sequences were used to maintain temporal continuity and use contextual information. However, for model inference, predictions were made for every frame in the input sequence ensuring a fine-grained resolution.

In Task 1 the classification model generated predictions for the central frame of each input window of training data. Therefore, during inference, test sequences were processed using a sliding window with a step size of 1 to ensure that every frame received a prediction. For consistency with the training procedure, the window size was set to 243 frames. An additional post-processing step was incorporated after the entire sequence for a subject was predicted. This step aimed to smooth out frame-level predictions by incorporating domain knowledge that each exercise typically lasts for at least 1 second, corresponding to 30 frames. Using this knowledge, if short sequences of frame-level predictions differed from the majority class within a window of size 30 frames, those frames were reassigned to the majority class of the window. This smoothing process reduced noise in the predictions and ensured more consistent and realistic classification outputs, aligning with the expected temporal dynamics of the exercises.

For the regression in Task 2, the architecture of VideoPose was designed to process the entire test sequence simultaneously, requiring no additional adjustments for inference [31]. The reported results were computed only for the pre-selected set of 3 axial and 6 lower body joints.

#### 2.3.2 Metrics

To evaluate the performance of the described models on the two tasks, the following metrics were used:

##### 1) Exercise Recognition

Accuracy and segment-wise F1-score at *k* (F1@*k*) were used to evaluate model performance on the classification task. Due to its frame-by-frame nature, exercise recognition resembles an action segmentation task which, as proposed by Lea et al. [61], is better evaluated using segment-wise F1 scores rather than sample-wise F1 score. This is because it penalizes over- and under-segmentation errors by measuring how well predicted segments match ground truth segments with at least *k*% temporal overlap. For each predicted segment, which is a sequence of identical labels, the Intersection over Union (IoU) with ground-truth segments of the same class is calculated as:

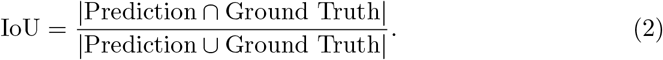

A predicted segment is considered a true positive (TP) if it overlaps with a ground-truth segment of the same class with an IoU *≥ k*. Predicted segments without a valid match are counted as false positives (FP), while unmatched ground-truth segments are false negatives (FN). Using these, Segment-F1@*k* is computed as:

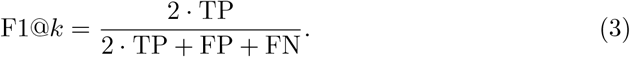

Following standard practice [61], we report results at Segment-F1@50 using a threshold of *k* = 0.5.

##### 2) Joint Angle Prediction

The Mean Per-Joint Angular Error (MPJAE) was used to evaluate the regression task. This metric, defined in Equation 1, calculates the mean angular difference in degrees between the predicted and ground truth angles for all relevant joints. It provides a robust measure of the model’s accuracy in predicting joint angles by capturing the average deviation across all frames and joints in the test sequence. Additionally, we report the Pearson correlation coefficient between the estimated and true angles as this is a common regression metric.

#### 2.3.3 Exercise performance metrics (EPMs)

To evaluate the performance of each physical exercise performed by the subjects, we calculated a set of EPMs. The EPMs were computed using both model predictions and ground truth labels, and their reliability was assessed by calculating the Pearson correlation coefficient between the two. We chose to compute the EPMs of the fine-grained sub-exercise actions rather than the complete exercises (Table 1), as this level of detail can offer important insights into an older adult’s performance. For example, taking longer to perform a lateral movement with the left leg compared to the right may suggest difficulties specific to one of the legs. Such distinctions can help identify which parts of an exercise are more physically challenging and may benefit from targeted intervention.

##### Total exercise duration

The duration of each exercise was computed in terms of the cumulative time each subject spent performing a specific sub-exercise action. This was calculated by counting the total number of frames corresponding to each sub-exercise action label and converting it to minutes based on the video frame rate.

Let *N*_frames,*e*_ represent the number of frames labeled as sub-exercise action *e*. The total duration in minutes of sub-exercise action *e*, denoted as *D*_*e*_, is calculated as:

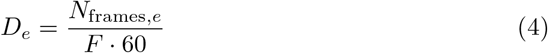

where *F* is the video frame rate (30 frames per second).

##### Number of repetitions

The number of repetitions for each sub-exercise action was calculated by identifying transitions in the frame-by-frame action labels. A new repetition was counted whenever the label transitioned from one sub-exercise action to another. To ensure robust counting, domain knowledge was incorporated to smooth out the label transition events. Specifically, a transition was only counted as a new repetition if the current label was the majority label in a window of 30 subsequent frames, corresponding to 1 second of activity. Formally, let *L*_*t*_ represent the label at frame *t*. A transition from frame *t* to frame *t* + 1 contributes to the repetition count *R*_*e*_ for exercise *e* if:

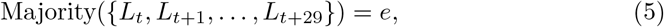

where Majority denotes the most frequent label within the 30-frame window. This smoothing process reduces noise in the label transitions and ensures only valid repetitions are counted.

##### Motion variability

Motion variability was included in this study as an important indicator of movement quality, neuromuscular control, and fall risk. Prior research has shown that increased variability in joint kinematics may reflect underlying stability impairments, particularly among older adults [46]. For example, high variability in hip flexion during tasks such as sitting, standing, or bending has been associated with reduced postural control and increased fall risk [62].

To quantify motion variability, we calculated the mean absolute deviation (MAD) of the rotational joint angles along the 3 degrees of freedom (i.e. lateral bending (*x*), flexion/extension (*y*), and axial bending (*z*)) during each exercise repetition. For a given joint *j*, angle *a ∈ {x, y, z}*, and exercise repetition *r* consisting of *n* time steps, MAD was defined as:

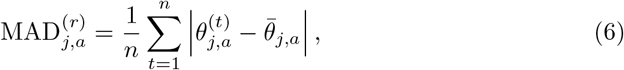

where 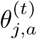 is the joint angle at time step *t* for joint *j* along rotational axis *a*, and 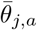 is the mean angle over the repetition. The time steps belonging to each exercise repetition were identified using the ground truth sub-exercise action labels from Task 1.

##### Range of motion (ROM)

ROM was included as one of our EPMs due to its well-established relationship with age-related mobility decline and reduced physical function in older adults [63]. In particular, this loss in ROM has been shown to be joint-specific, likely reflecting the varying patterns of joint usage accumulated over a lifetime. Such joint-level insights can inform the personalization of exercise routines for older adults. In our study, ROM was defined as the difference between the maximum and minimum joint angle values observed within each exercise repetition. For a given joint *j*, rotational angle *a ∈ {x, y, z}*, and exercise repetition *r* consisting of *n* time steps, the ROM was computed as:

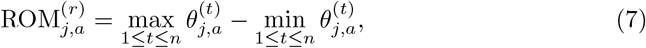

where 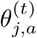 represents the joint angle at time step *t* for joint *j* along rotational axis *a*.

## 3 Results

This section begins with a description of the data characteristics, followed by an overview of the performance of the implemented DL model on (1) exercise recognition and (2) joint angle prediction. Finally, the accuracy of EMPs estimated from model predictions is compared to those derived from ground truth data.

### 3.1 Dataset

The study included seven older adult participants, consisting of four males and three females, aged between 65 and 94 years (mean age: 82.4 ± 8.1 years). A summary of their demographic characteristics is presented in Table 2.

**Table 2:** Demographic characteristics. Overview of the height, foot length, age, gender, and body weight at the time of data collection for each subject.

| Subject ID | Height (cm) | Foot Length (cm) | Age range (years) | Gender | Weight (kg) |
| --- | --- | --- | --- | --- | --- |
| 1 | 168 | 27.0 | 80-84 | Female | 111.8 |
| 2 | 170 | 26.5 | 65-69 | Male | 79.8 |
| 3 | 165 | 26.0 | 85-89 | Female | 52.2 |
| 4 | 170 | 26.5 | 85-89 | Male | 52.0 |
| 5 | 175 | 28.0 | 80-84 | Male | 91.6 |
| 6 | 163 | 25.5 | 90-94 | Female | 66.0 |
| 7 | 178 | 26.5 | 75-79 | Male | 70.0 |

Table 3 provides an overview of the collected exercise data that was used in the analysis of this study. Each participant completed between 5 and 8 measurement sessions, resulting in a total duration ranging from 67 to 94 minutes per subject. The mean duration per session varied between 10 and 15 minutes which reflects differences in exercises pace and individual performance across subjects.

**Table 3:** Dataset characteristics. Overview of the data collected for each subject including the number of measurement trials, the total duration of all measurements in minutes and the mean duration of a measurement in minutes.

| Subject | Measurements | Total Duration (min) | Mean Duration (min) |
| --- | --- | --- | --- |
| S1 | 5 | 67.33 | 13.47 |
| S2 | 6 | 94.14 | 15.69 |
| S3 | 6 | 90.72 | 15.12 |
| S4 | 6 | 94.63 | 15.77 |
| S5 | 5 | 71.98 | 14.40 |
| S6 | 7 | 87.61 | 12.52 |
| S7 | 8 | 82.53 | 10.32 |

### 3.2 DL Model performance

We evaluated the performance of the DL architecture on both tasks: exercise recognition (Task 1) and joint angle estimation (Task 2). The results are summarized in Table 4 in terms of the mean LOSO CV performance metrics. To better understand the strengths and limitations of the model, we provide a detailed analysis of the performance in each task in the upcoming sections.

**Table 4:** LOSO CV performance of the DL model on Task 1 (Exercise recognition) and Task 2 (joint angle estimation).

| Task | Metric | LOSO CV mean (± SD) |
| --- | --- | --- |
| Exercise recognition | F1@50 | $0.92 \pm 0.04$ |
| | Accuracy | $0.88 \pm 0.04$ |
| Joint angle estimation | MPJAE [$^{\circ}$] | $7.7 \pm 0.91$ |
| | Correlation (r) | $0.75 \pm 0.05$ |

#### 3.2.1 Exercise recognition (Task 1)

From Figure 3 it can be seen that the model performed quite consistently in recognizing the different sub-exercise actions. The observed outliers for the last exercise of touching a cup with each foot, comprising the last four sub-exercise actions, are due to one subject (Subject 2). During data collection this participant experienced particular difficulties with the exercises, exhibited restricted mobility, and required assistance during some of the movements. Additionally, the model struggled to classify random movements, likely due to their inherently higher motion variability.

**Fig. 3:**
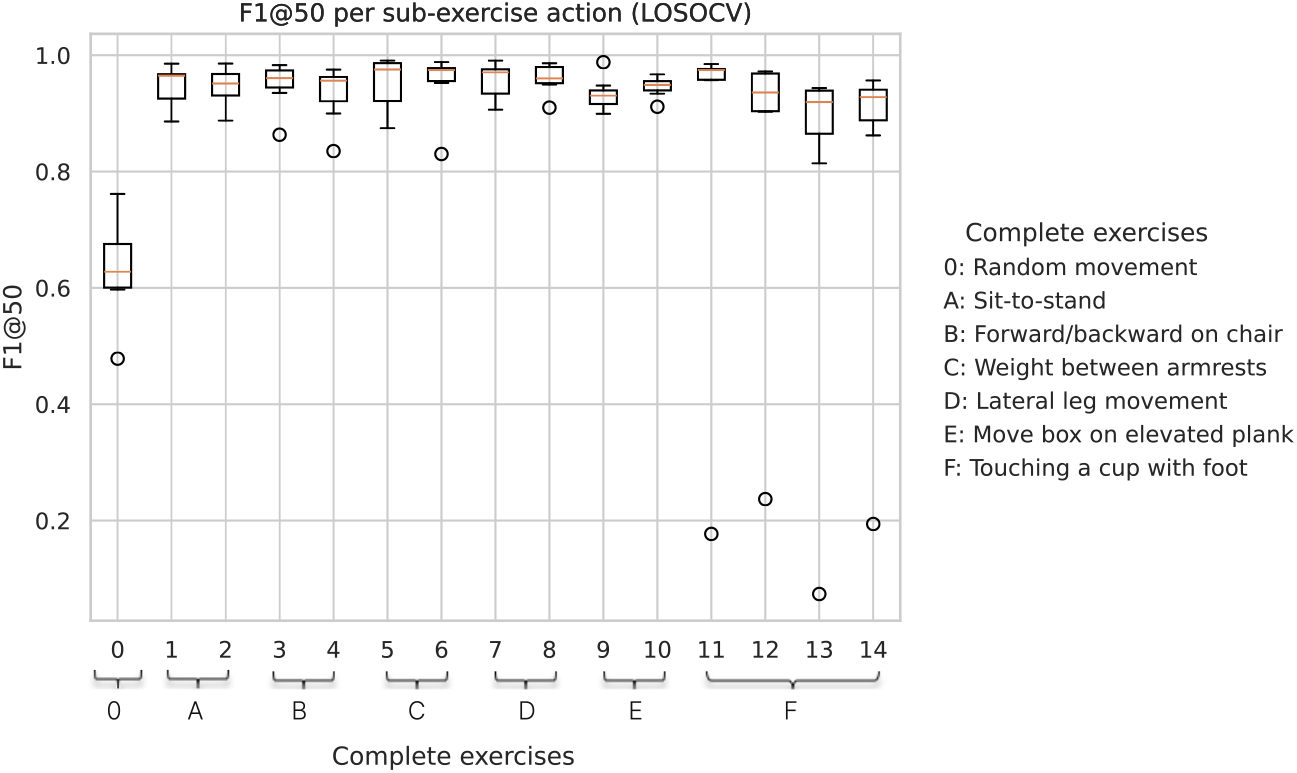
Model performance on exercise recognition (LOSO CV). Each boxplot represents the F1@50 scores per sub-exercise action for all subjects. For clarity the sub-exercise actions are grouped into their corresponding complete exercises (see Table 1 for detailed description on the annotations)

Furthermore, the model output example of annotated time frames in Figure 4 illustrates that the model performed well in detecting the action boundaries in frame-by-frame annotation when compared to the manually annotated ground truth. However, it also highlights the challenge of accurately identifying the random movements that occur between exercises, likely because this label describes a wide variety of movement types. In this particular case, the subject may have been adjusting their position on the chair before initiating the next exercise, which the model interpreted as the bending backward action. Despite this, the overall classification of the complete exercise remains correct, as the model successfully identifies the transition to the bending forward/backward exercise sequence.

**Fig. 4:**
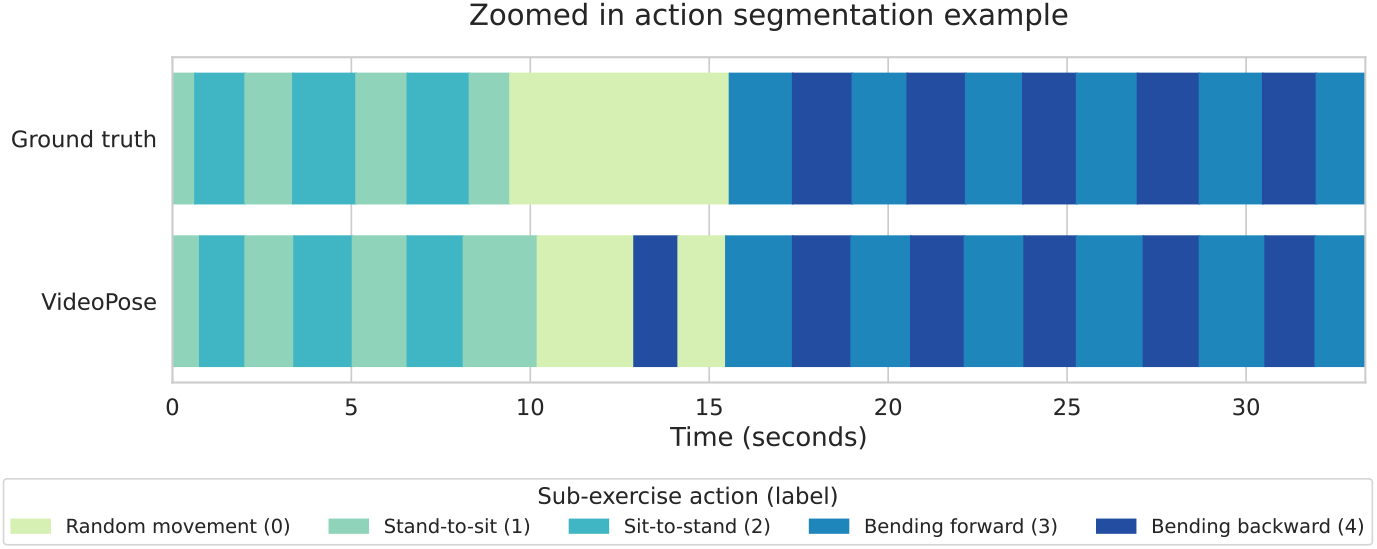
An example time segment of a subject trial with action segmentation. The figure displays sub-exercise action labels over time, comparing the manually annotated ground truth with predictions from our DL model. This example is from a transition where the subject switched from performing the sit-to-stand exercise to the forward and backward bending movement on a chair.

#### 3.2.2 Joint angle estimation (Task 2)

Figure 5 presents the LOSO CV results for estimating the three rotational angles across the 9 selected joints (3 axial and 6 lower body). Overall, the model demonstrates strong performance, though errors vary by joint. Notably, the hip and knee joints exhibit the highest estimation errors. This aligns with previous findings that highlighted greater variability and complexity in predicting lower body joint angles due to increased mobility and degrees of freedom [64].

**Fig. 5:**
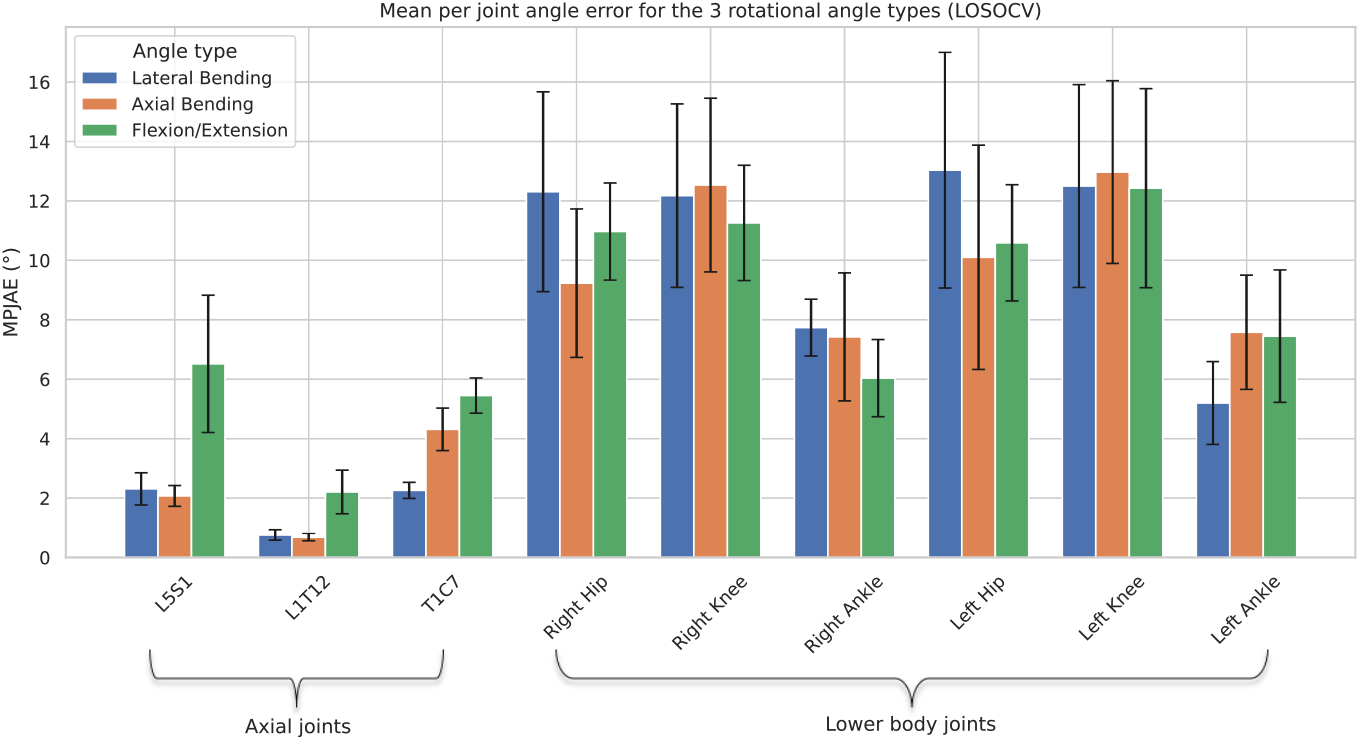
Model performance on joint angle prediction (LOSO CV). Each bar represents the mean MPJAE in degrees over all subjects and the error bars show the variability between LOSO CV folds. For each of the pre-selected 9 joints, the three rotational angles estimation errors are shown.

It is important to interpret these results within the context of the specific exercises, as the estimation errors differ depending on which joint is involved and how it contributes to the movement. To illustrate this in terms of the continuous joint angle signals generated by the model, Figure 6 presents a frame-wise example of an exercise sequence. In the first segment, where the subject is moving a weight between the armrests while seated, the hip flexion angle is not accurately estimated. However, this inaccuracy is less concerning, as hip flexion is not functionally relevant for this particular exercise. In contrast, the axial rotation of the L1T12 joint which is located along the thoracic-lumbar spine and is more engaged during this movement, is well estimated. Furthermore, in the subsequent sit-to-stand exercise, where hip flexion plays a central role in assessing movement quality and stability, the predicted signal aligns closely with the ground truth. This indicates strong performance of the model in estimating clinically relevant joint angles when interpreted in the context of the exercise.

**Fig. 6:**
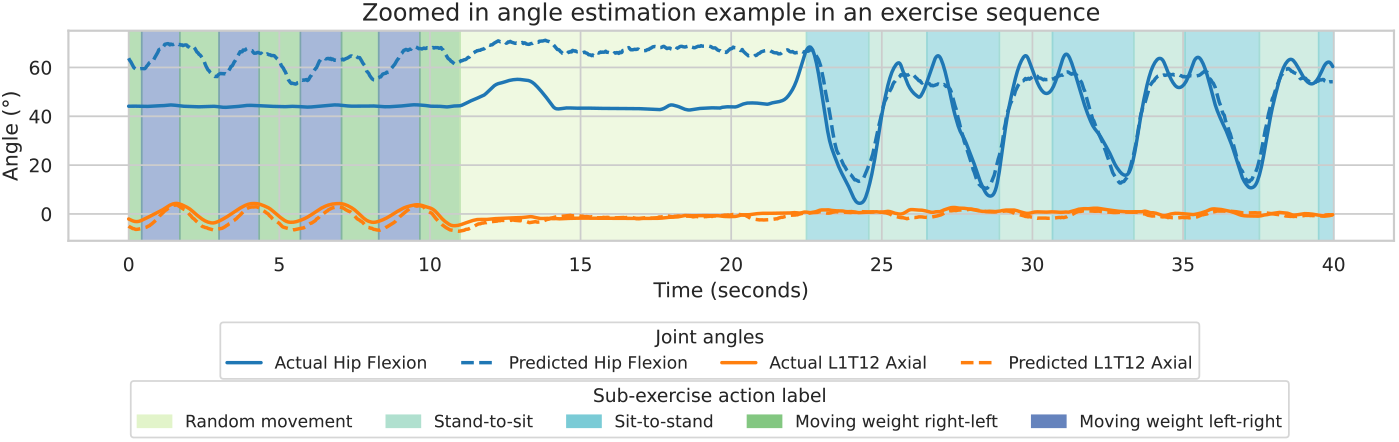
An example segment of the predicted and ground truth hip flexion and L1T12 axial rotation angles during the transition between two exercises. On the background are the sub-exercise action annotations illustrated with different colors.

### 3.3 Exercise performance metrics estimated from model predictions

The first two EPMs, total exercise duration and number of repetitions, were derived from the frame-level sub-exercise action labels in Task 1. As shown in Figure 7, the predicted exercise duration closely matches the ground truth, with a strong correlation coefficient of r = 0.93 with a 95% confidence interval (CI) [0.90, 0.95]. The deviations from the true values indicate that the model is more suitable for broader time estimations, such as on the scale of minutes, rather than highly precise measurements in the millisecond range. Similarly, the predicted repetition count strongly aligns with the true values, achieving a correlation of r = 0.86 with CI [0.80, 0.90]. Overall, the performance is consistent across exercises and subjects, with the exception of Subject 2, who had drops in action recognition performance, as previously detailed in Section 3.2.1.

**Fig. 7:**
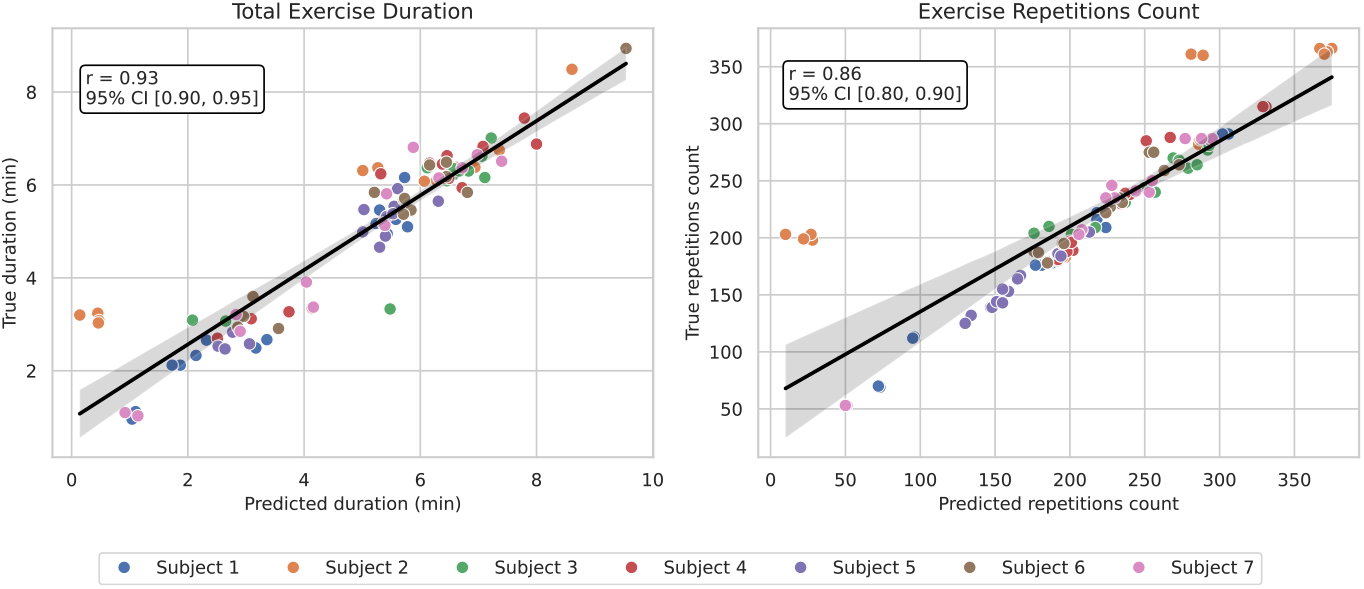
Pearson’s correlation coefficient (*r*) between predicted and actual exercise duration (left) and repetition count (right) across subjects. Each point represents a sub-exercise action, color-coded by subject. The solid black line indicates the linear regression fit, and the shaded area shows the 95% confidence interval.

The other two EPMs, motion variability (in terms of MAD) and ROM, were computed using the joint angle estimations from Task 2. As mentioned in the Task 2 results we expected to observe varying results in the calculation of these EPMs depending on the context in terms of joint, angle type and exercise. An example of the variability in results can be seen in Figure 8, where only the flexion angles are used to calculate MAD and ROM values. Strong correlations (up to 0.9) are observed for the hips and knees across multiple exercises. This is especially relevant for the sit-to-stand exercise (labels 1 and 2), which is among the most physically demanding ones and poses the highest stability risk for this population. In contrast, the ankles and the three axial joints (L5S1, L1T12, T1C7) consistently show lower performance in both MAD and ROM for the flexion/extension angle. This is likely because the corresponding exercises did not involve significant flexion in those joints, making their motion patterns subtler and their estimations noisier, which led to noisier calculations of EMPs.

**Fig. 8:**
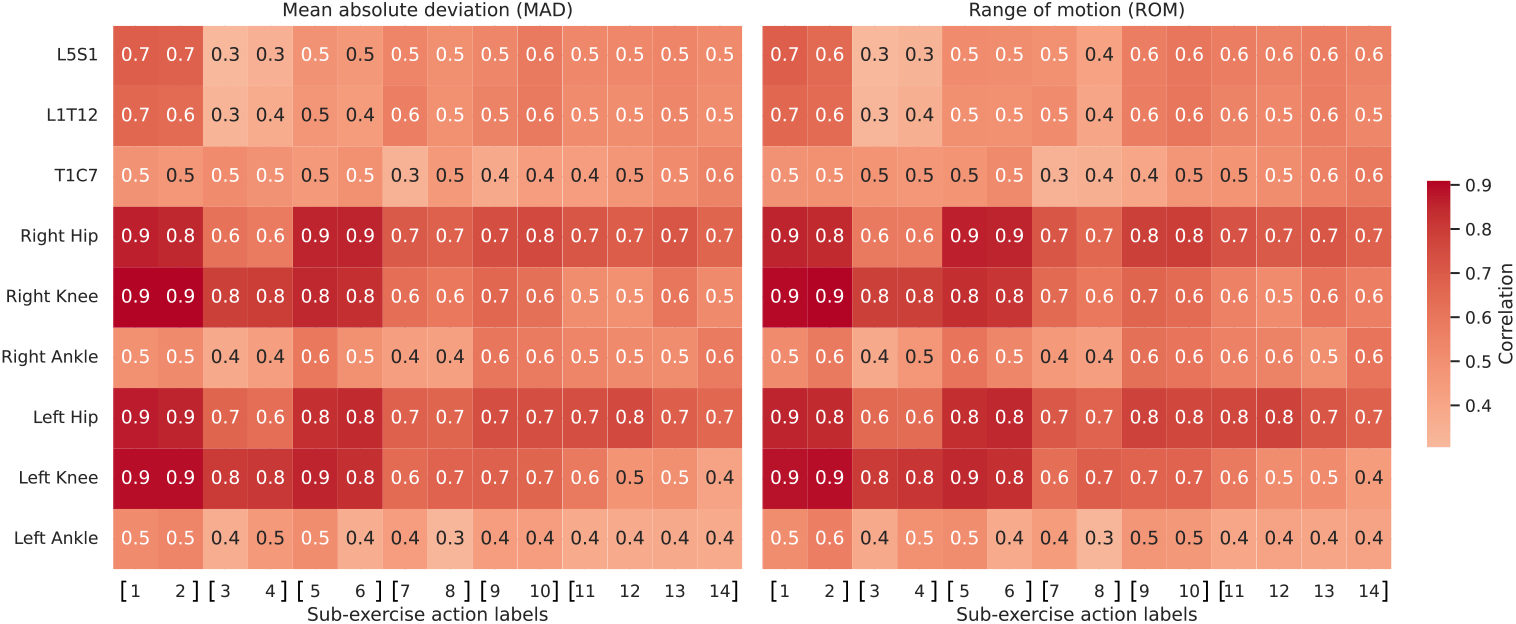
Correlation between the predicted and true mean absolute deviation (MAD, left) and range of motion (ROM, right) for the flexion/extension angle. Each cell represents the correlation for a specific joint (rows) and sub-exercise action (columns), averaged over all subjects. The action labels are grouped to their corresponding complete exercises using square brackets ‘[]’. The varying scores highlight the influence of joint function and exercise type on model performance.

Our primary objective in computing these ETMs is to demonstrate their potential in supporting clinicians to monitor exercise performance in older adults over time. Figure 9 illustrates this by showing the evolution of MAD and ROM values for the hip and knee flexion angles across 8 repetitions of the sit-to-stand exercise for a single participant. In this example, the participant displays an overall stable hip and knee flexion movement, as reflected by relatively consistent MAD values. However, during two early repetitions (circled in red), both hip and knee ROM values are noticeably lower, indicating the subject did not fully extend their joints when standing up. The reduced movement is also accompanied by low MAD scores, which does not seem to be due to improved movement variability, but rather due to limited ROM. This highlights the importance of interpreting MAD alongside ROM to avoid misleading conclusions. For instance, a consistent decline in both ROM and MAD over time might suggest that the participant is no longer exercising using their full motor capabilities. This could be either due to emerging physical pain or due to a lack of motivation. In contrast a low ROM coupled with high MAD values might indicate stability or balance issues. Identifying such patterns can facilitate an intervention by the caregivers allowing them to adapt the exercise program to better suit the participant’s evolving needs.

**Fig. 9:**
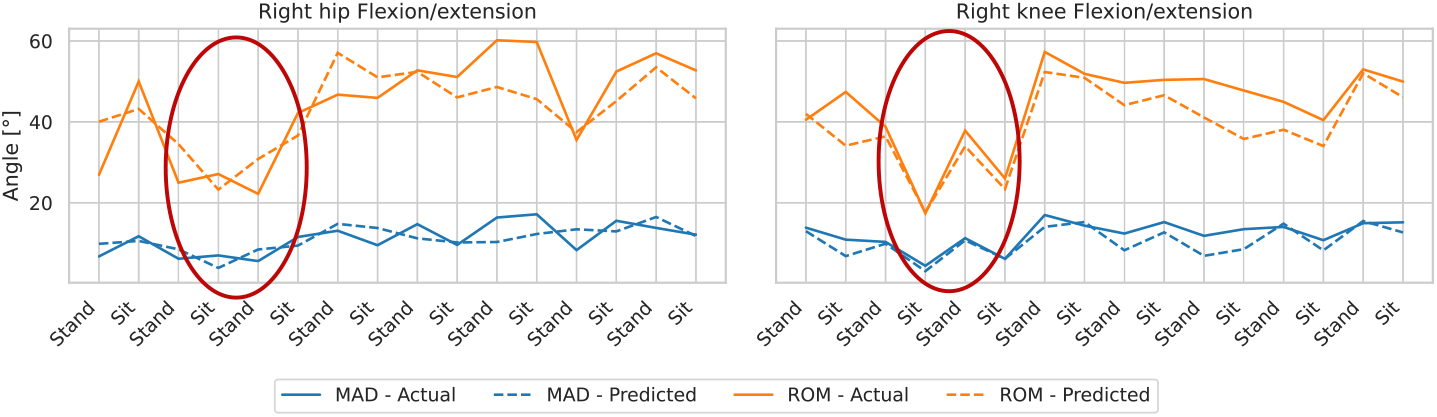
Example segment of the sit-to-stand exercise displaying the predicted and actual MAD (blue) and ROM (orange) values for the flexion/extension angle of the right hip (left) and ankle (right). The figure shows the changes in both ETMs over time during each sub-exercise action of 8 repetitions of the complete sit-to-stand exercise. The part highlighted with a red eclipse is a short segment where the participant did not stand up as high as in other repetitions, but quickly corrected themselves.

## 4 Discussion

This study presents a novel method for the unsupervised monitoring of older adults’ exercise performance using a single-camera system combined with deep learning. We demonstrated that a DL model can be trained to successfully recognize exercise actions and estimate joint angles in older adults. Our implementation relied on existing tools: AlphaPose for 2D pose estimation and VideoPose for processing temporal pose data for both classification and regression tasks. While the system showed promising performance, future work could explore whether alternative 2D pose estimators, such as OpenPose [65] and PoseNet [66], or different model architectures might improve accuracy. However, it is important that any future improvements take into account the computational power required to run these algorithms to ensure that their deployment in real-world scenarios is practical.

A key contribution of this study is the use of 3D joint angles for exercise monitoring, rather than the commonly used 3D joint coordinates. By focusing on angles, our approach offers a more interpretable and task-relevant measure of joint movement, particularly for assessing outcomes like motion variability and range of motion, highlighting their value for monitoring older adults’ exercises. The upper body joints were excluded from the analysis due to difficulties in data collection, such as inconsistent calibration of the upper body skeleton, which introduced noise into the motion data. This limitation highlights the challenges of capturing precise motion data in older adults, particularly for exercises involving subtle or limited joint movements. Another potential concern could be our use of motion sensors as a ground truth for joint angles, which are known to have a degree of measurement inaccuracy [67]. However, this is not a major issue for our application, which focuses on tracking changes over time rather than clinical evaluation of individual exercise execution. Even with minor sensor inaccuracies, longitudinal trends in ROM and motion variability still provide meaningful insights into functional progression.

Additionally, this work focused on a few key exercise outcomes, but more metrics, such as limb coordination or muscle power, could be incorporated in future studies. Our system offers significant potential for longitudinal monitoring of exercise performance due to the contactless nature of the data. This could enable caretakers in residential homes to adjust exercise programs to better fit the individual needs of older adults. The approach aligns with the WHO’s recommendations for healthy aging, which emphasize the importance of maintaining functional ability through regular physical activity and personalized exercise interventions [23]. The framework could also be adapted for real-time processing to provide corrective feedback during exercises, further improving exercise quality and safety [68]. Importantly, the method underscores the potential of our system to provide meaningful insights into exercise performance in accordance with established guidelines for healthy aging. This method offers a cost-effective solution for care homes to facilitate regular exercise sessions for residents without requiring active caregiver supervision. Staffing is one of the primary costs associated with care homes in Europe [69], and this approach could help minimizing labor costs by utilizing a simple webcam with no high-resolution requirements, reducing the need for expensive equipment or additional personnel.

A limitation of this study is the small sample size and the fact that data collection was conducted in a single facility, which may restrict the generalization of the findings. However, the sample captured the heterogeneity within this population, including varying levels of frailty and differences in movement patterns. The controlled environment of the facility, with minimal visual distractions such as paintings, plants, or other objects, may have facilitated the accuracy of the motion tracking system. Future studies should explore whether the approach remains effective in more visually complex environments, including home settings, where factors like furniture, lighting conditions, and background movement could impact tracking performance. Finally, our current approach is limited to single-person analysis. Extending the system to group exercise settings could enhance its applicability by enabling interactive sessions for older adults, fostering engagement and social interaction during physical activity [70].

## 5 Conclusion

This study aimed to address the need for scalable and cost-effective solutions for unsupervised monitoring of older adults’ exercise performance. By leveraging an existing model acrhitecture for 3D pose estimation (VideoPose) we successfully adapted it to recognize exercises and estimate rotational angles for axial and lower body joints, enabling the assessment of key exercise performance metrics such as duration, repetition count, motion variability, and range of motion.

This work highlights the potential of a deep learning-based system to monitor exercise performance in older adults, offering opportunities for longitudinal tracking to support tailored physical activity programs. Future improvements, such as enabling real-time processing and extending the system to group exercise analysis, could further enhance its utility in promoting the health and independence of older adults.

## Supplementary information

No supplementary materials are provided for this study.

## Acknowledgments

We thank all the participants who generously gave their time and effort to contribute to this study as well as the Flanders AI Research Program for supporting the project.

## Declarations

### Ethics approval and consent to participate

All participants signed an informed consent form. The study was approved by the ethical committee of KU Leuven and the university hospital UZ Leuven (S61015).

### Consent for publication

Not applicable.

### Data availability

The datasets generated during and/or analyzed during the current study are not publicly available due to privacy concerns.

### Materials availability

All materials used in this study are commercially available.

### Code availability

The code used for analysis in this study is not publicly available but is available from the corresponding author upon reasonable request.

### Author contribution

Conceptualization: Vayalet Stefanova, Benjamin Filtjens Bart Vanrumste; Methodology: Vayalet Stefanova, Benjamin Filtjens, Bart Vanrumste; Software: Vayalet Stefanova, Benjamin Filtjens; Validation: Vayalet Stefanova; Data Curation: Evelien Maeyens, Jeroen Brughmans, Karen Gilis, Dimitri Vargemidis; Writing – Original Draft: Vayalet Stefanova; Writing – Review and Editing: Julien Lebleu, Leen Stulens, Karen Gilis, Sanne Broeder, Mieke Deschodt, Benedicte Vanwanseele, David Beckwée, Moran Gilat, Benjamin Filtjens, Bart Vanrumste; Funding Acquisition: Benjamin Filtjens, Bart Vanrumste.

